# Scalable Approach to Medical Wearable Post-Market Surveillance

**DOI:** 10.1101/2023.11.14.23298488

**Authors:** Richard M. Yoo, Ben T. Viggiano, Krishna N. Pundi, Jason A. Fries, Aydin Zahedivash, Tanya Podchiyska, Natasha Din, Nigam H. Shah

## Abstract

**Objective:** We sought to develop a weak supervision-based approach to demonstrate feasibility of post-market surveillance of wearable devices that render AF pre-diagnosis.

**Materials and Methods:** Two approaches were evaluated to reduce clinical note labeling overhead for creating a training set for a classifier: one using programmatic codes, and the other using prompts to large language models (LLMs). Probabilistically labeled notes were then used to fine-tune a classifier, which identified patients with AF pre-diagnosis mentions in a note. A retrospective cohort study was conducted, where the baseline characteristics and subsequent care patterns of patients identified by the classifier were compared against those who did not receive pre-diagnosis.

**Results:** Label model derived from prompt-based labeling heuristics using LLMs (precision = 0.67, recall = 0.83, F1 = 0.74) nearly achieved the performance of code-based heuristics (precision = 0.84, recall = 0.72, F1 = 0.77), while cutting down the cost to create a labeled training set. The classifier learned on the labeled notes accurately identified patients with AF pre-diagnosis (precision = 0.85, recall = 0.81, F1 = 0.83). Those patients who received pre-diagnosis exhibited different demographic and comorbidity characteristics, and were enriched for anticoagulation and eventual diagnosis of AF. At the index diagnosis, existence of pre-diagnosis did not stratify patients on clinical characteristics, but did correlate with anticoagulant prescription.

**Discussion and Conclusion:** Our work establishes the feasibility of an EHR-based surveillance system for wearable devices that render AF pre-diagnosis. Further work is necessary to generalize these findings for patient populations at other sites.

## BACKGROUND

With the advent of consumer-facing devices such as Apple Watch[1] and FitBit[1,2] that can render *pre-diagnosis* such as atrial fibrillation (AF) based on collected photoplethysmography and electrocardiogram (ECG) data, medical wearables now have the potential to affect diagnosis rates and initiate cascades of medical care[3,4]. While these devices undergo pre-market validation to obtain FDA clearance[5], there remains limited information on their post-market use and clinical utility.

One of the primary challenges in *post-market surveillance* for wearables is that current medical terminologies do not have terms for representing wearable use. For example, much of outcomes research is built around medical diagnosis codes used for billing purposes, such as the International Classification of Disease (ICD)[6] or Current Procedural Terminology (CPT)[7], neither of which contain terms for describing wearable use. Therefore, no structured data exists in electronic health records (EHRs), and unstructured data such as clinical notes must be parsed to obtain the information.

Advances in deep learning-based natural language processing (NLP) methods[8–10], in particular their application to clinical note classification tasks[11,12], have shown to outperform traditional pattern and rule-based approaches. However, these deep-learning based classifiers require large, hand-labeled training sets that can be costly and time-consuming to generate. For a post-market surveillance strategy based on EHRs to be widely implemented, a scalable approach is necessary to reduce the labeling overhead. In particular, pre-trained large language models (LLMs) have the potential to dramatically accelerate this process through prompting, but their application to the medical domain has not been clear.

## OBJECTIVE

We developed a weak supervision-based approach to demonstrate feasibility of post-market surveillance of wearable devices that render AF pre-diagnosis, and evaluated its efficacy. The first aim of this study was to evaluate two approaches for generating labeling heuristics for training sets: one using programmatic codes, and the other using prompts submitted to LLMs. A *label model* derived from such labeling heuristics probabilistically assigns labels to clinical notes, about whether the note contains a mention of the patient receiving AF pre-diagnosis from a wearable device. The second aim was to use a training set labeled by the label model to fine-tune a *classifier* that identifies mentions of AF pre-diagnosis in a clinical note, and to evaluate its performance. Our third aim was to summarize the clinical characteristics of patients identified by the classifier, and compare them to patients who were not alerted to AF pre-diagnosis.

## MATERIALS AND METHODS

### Cohort Identification

We used the Stanford Medicine Research Data Repository (STARR)[13], which contains EHR-derived medical records from the inpatient, outpatient, and ER visits at Stanford Health Care and the Lucile Packard Children’s Hospital. Methods for dataset creation have been previously described[13]. We queried STARR to retrieve all clinical notes that contain a mention of a wearable device (see Supplemental Table 1 for the complete list of search terms), resulting in 86,260 notes from 34,329 unique patients. To match the FDA guidance for pertinent cardiovascular algorithms[5], we excluded patients less than 22 years of age when the note was written, leaving 78,323 notes from 30,133 unique patients. We further limited to notes written on or after January 1, 2019, since the first publicly available AF detection feature became available in December 2018[14]. The resulting cohort comprised 56,924 clinical notes from 21,332 unique individuals.

### Manual Labeling Process

Among the 56,924 clinical notes with mentions of wearables, we manually labeled 600 notes to construct a test set to assess the performance of the label model and the classifier. Specifically, to remove duplicate texts and cover as many patients as possible, we randomly selected 600 unique patients, then selected one note for each patient. These notes were then labeled independently by two data scientists, and differences were adjudicated by two physicians. A clinical note was labeled as positive when it was clear that the patient received automated AF notification from the wearable, or that the patient initiated an on-demand measurement (e.g., ECG strip) which resulted in AF pre-diagnosis.

In addition to the test set, to aid in label model development, we prepared another set of 600 notes that was used as the development set. This set was manually labeled by a single data scientist, using a labeling guideline (see Supplemental Material 1) that was developed as part of the test set generation.

### Label Model Derivation 1: Code-Based Labeling Functions

We then derived a label model that used a weak supervision-based approach to programmatically generate training set labels. Specifically, as shown in Figure 1(a), we used data programming[16], where labeling heuristics are expressed as *labeling functions* (LFs) that encode domain insights. In particular, *code-based* labeling functions (see Figure 1(a), Approach 1) capture heuristics using programmatic language constructs. Predictions from these labeling functions are then combined to learn a generative *label model*.

**Figure 1.**
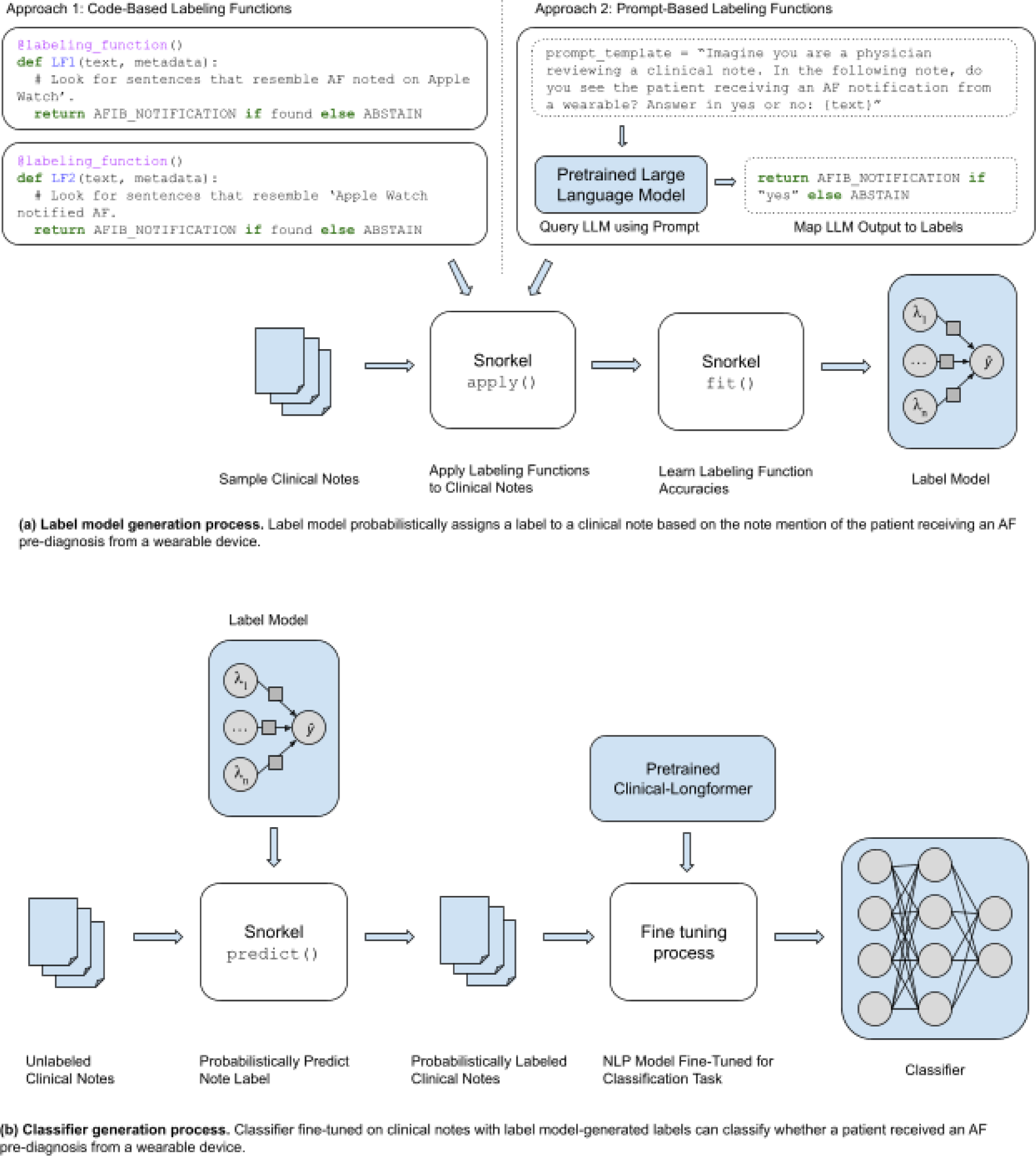
Label model and classifier generation process using the Snorkel[15] framework. Figure 1(a) describes the label model generation process, where labeling heuristics are expressed as either code-based or prompt-based labeling functions. Snorkel then applies the labeling functions to the sample clinical notes and fits a generative model on the labeling function predictions. In Figure 1(b), the obtained label model is used to probabilistically assign labels for a large number of unlabeled clinical notes, which are then used to fine-tune a classifier to detect whether a patient received AF pre-diagnosis from a wearable device.

We used the Snorkel framework[15] to implement data programming. A preprocessing framework[17] was applied to preprocess our notes, where the clinical notes were split into sentences using the spaCy[18] framework, with a specialized tokenizer to recognize abbreviations and terms specific to medical literature. Thus parsed grammatical information was made available to the labeling functions as metadata to each sentence.

We then observed the notes in the development set to understand how AF notifications were described, and expressed each pattern as a labeling function using the grammar metadata, or simple dictionary lookup. The labeling function development process was iterative, where each labeling function was further optimized based on the performance metrics (i.e., precision, recall, and F1) calculated by the Snorkel framework on the development set.

Once developed, we ran the labeling functions on the samples, then instructed the Snorkel framework to fit a generative model on the output. We used 10-fold cross validation on the test set, and chose the best performing label model. This model was then run across the entire 56,924 clinical notes to probabilistically assign their labels.

### Label Model Derivation 2: Prompt-Based Labeling Functions

We then derived another label model that applies *prompting* techniques to query LLMs (see Figure 1(a), Approach 2). This approach leverages the observation that a pre-trained LLM can generalize to a new task that it was not trained for[19]. Specifically, inside a labeling function, a clinical note is presented to the LLM as part of a *prompt*, which queries the LLM for certain characteristics of the note that would be relevant to determining the label. The LLM’s answer to the prompts are then mapped to predicted labels returned by a labeling function.

Compared to code-based labeling functions where heuristics to determine note labels are expressed using software language constructs, prompt-based labeling functions accept *natural language prompts* to query the characteristics of clinical notes. This approach has been shown to provide remarkable performance gains while reducing overall cost (i.e., engineering time, expert face time) to author and optimize labeling functions[20].

Similar to how code-based labeling functions were developed, different natural language prompts were tried iteratively, while using Snorkel to assess performance. We tried two different *open* LLMs: T0++[21] and Flan-T5-XXL[22]. T0++ is an LLM that is trained specifically with multiple prompts for multiple tasks, which often shows better performance than a much larger model like GPT-3 on prompting tasks. Flan-T5-XXL also leverages similar multitask training. Both models are similar in their size (11 billion parameters), but were released about a year apart, reflecting the advances in machine learning techniques. We could not leverage *closed* LLMs such as GPT and ChatGPT, since at the time of this research they lacked a HIPAA compliant API. We used Manifest[23] to set up the Hugging Face releases of each LLM in a HIPAA-compliant secure computing environment.

To properly assess the tradeoff that the prompt-based approach introduces between label model performance and development cost, we time-boxed ourselves to a single day of prompt-based labeling function development, and observed how much of the performance with code-based labeling functions could be attained. In comparison, we devoted an unbounded amount of time for code-based labeling function development and optimization—amounting to two weeks of data scientist time and three days of clinician time. The same 10-fold cross validation on the test set was used to evaluate the label model. To remediate LLM performance variances across runs, we repeated the same experiment 3 times, and observed the average performance.

### Classifier Development

Clinical notes that were probabilistically labeled by the label model were then used to fine-tune a large, NLP-based classifier, Clinical-Longformer[12] (see Figure 1(b)). The resulting classifier takes plain note text as the input, and classifies the note as positive (i.e., includes mention of a patient receiving an AF notification, or a patient-initiated on-demand cardiac testing/ECG resulting in AF pre-diagnosis) or negative. When a classifier is trained on the label model output, it enables generalizing beyond the labeling heuristics that were encoded in the labeling functions themselves, such that the classifier can recognize more patterns than those that were expressed in labeling functions.

For each clinical note in the training set we formed a pair of note text and its label, and presented a sequence of those pairs as the input to the pretrained Clinical-Longformer while fine-tuning it for a sequence classification task. Clinical-Longformer uses sub-word tokenization, and 94% of our notes analyzed had fewer tokens than the Clinical-Longformer’s maximum input length of 4,096 tokens. Notes with more tokens were trimmed.

We fine-tuned multiple classifiers with varying training set sizes, and used the test set to assess their performance. Specifically, for a single fine-tuning run, we took regular snapshots of the classifier and chose the snapshot with the best F1 score on the test set as the representative snapshot of the run. Adam optimizer was used, with learning rate ramping up to 1e^-5^ followed by linear decay over three epochs.

The test set was never presented to the model during the fine-tuning process. Since our dataset was highly skewed towards negative samples, we stratified the training set to maintain a 1:2 ratio between the positive and negative samples. All samples in the training set were chosen randomly. The classifier with the best F1 score was then run across the entire 56,924 clinical notes to identify all AF pre-diagnosis incidents.

### Retrospective Cohort Study

Using the pre-diagnosis mentions flagged by the classifier, we identified patients who received AF pre-diagnosis, and performed a retrospective descriptive cohort study comparing the characteristics of the patients who received pre-diagnosis to those who did not, utilizing the same STARR dataset.

First we considered all the patients in the cohort regardless of their prior AF diagnosis. We compared the demographics, CHA_2_DS_2_-VASc[24] score, and its related comorbidities on the date the index note was created. When a patient had received one or more AF pre-diagnosis from a wearable, we defined the oldest note with pre-diagnosis as the index note since it is the most likely to drive downstream medical intervention. When a patient had not received any pre-diagnosis, the oldest note with mention of a wearable was chosen as the index note.

We then focused on those patients who did not have prior AF diagnosis. A patient was filtered out if the patient had received AF diagnosis, defined as an ambulatory or inpatient encounter with SNOMED code 313217, prior to the index note. We then compared the same demographics and comorbidities between those who received pre-diagnosis and those who did not, on the date the index note was created.

Lastly, we further confined the analysis to the patients who received clinician AF diagnosis within 60 days of the index note. Same as before, we excluded patients who had prior AF diagnosis before the index note. Patients were then grouped based on whether they had received AF pre-diagnosis from a wearable, and characterized on the date they received index AF diagnosis. In addition to the demographics and comorbidities, we also compared warfarin and Direct Oral Anticoagulant (DOAC; i.e., apixaban, dabigatran, edoxaban, rivaroxaban) prescription rates between the two groups of patients.

### Statistical Analysis

Throughout the analysis, when compiling patient race information, we used the 5 categories of race defined by the U.S. Census, and cast hispanic as a dedicated ethnicity. 10% of the patients were missing race information, so we categorized them as belonging to *undisclosed* race. Five patients were missing sex information, and were not included in the analysis. The Stanford Institutional Review Board (Stanford, CA) approved this study.

For hypothesis testing, we used 1-tailed Welch’s *t* test for continuous variables, and chi-squared test for categorical variables. Statistical analysis was performed using Pandas[25] 1.3.0 and SciPy[26] 1.7.0, running on Python 3.9.6 configured through Conda 4.5.11.

## RESULTS

### Label Model Performance 1: Code-Based Labeling Functions

In total, eight code-based labeling functions were authored over the course of two weeks. Most labeling functions utilized the grammatical information present in the metadata, while one used simple dictionary-based lookup. Table 1 provides the performance of each labeling function, followed by the combined label model.

**Table 1.**
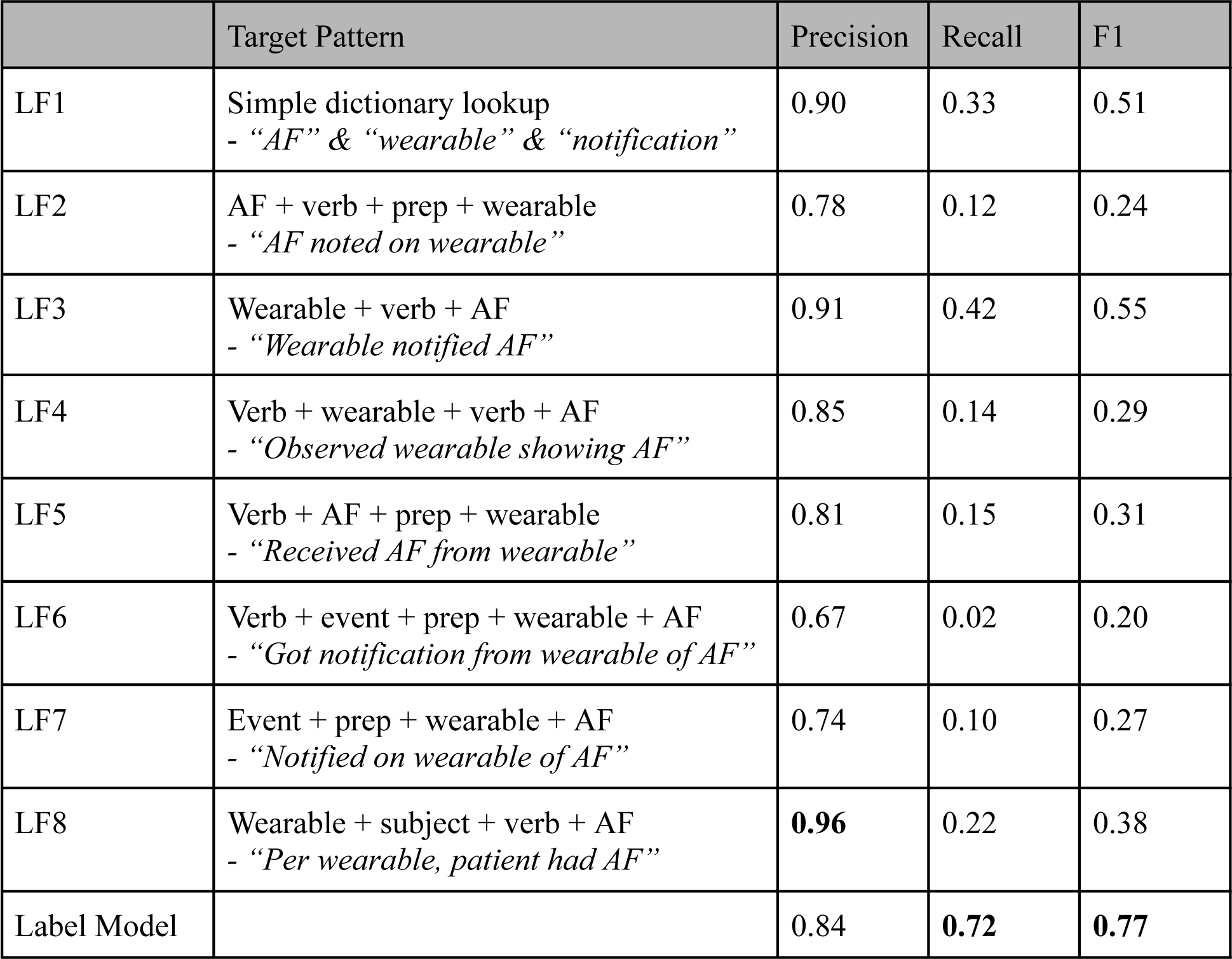
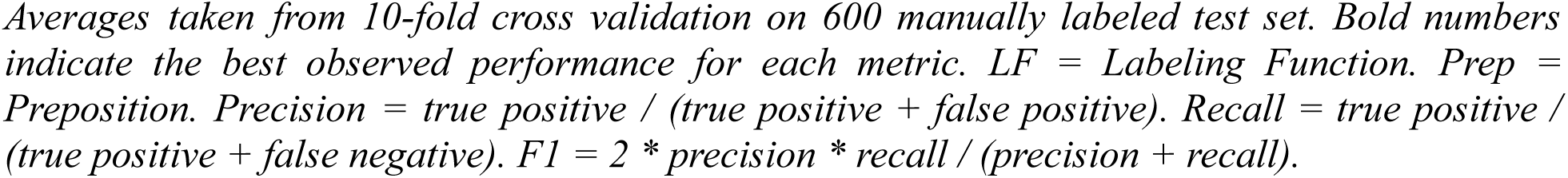
Label model performance with code-based labeling functions.

|  | Target Pattern | Precision | Recall | F1 |
| --- | --- | --- | --- | --- |
| LF1 | Simple dictionary lookup<br>- “AF” & “wearable” & “notification” | 0.90 | 0.33 | 0.51 |
| LF2 | AF + verb + prep + wearable<br>- “AF noted on wearable” | 0.78 | 0.12 | 0.24 |
| LF3 | Wearable + verb + AF<br>- “Wearable notified AF” | 0.91 | 0.42 | 0.55 |
| LF4 | Verb + wearable + verb + AF<br>- “Observed wearable showing AF” | 0.85 | 0.14 | 0.29 |
| LF5 | Verb + AF + prep + wearable<br>- “Received AF from wearable” | 0.81 | 0.15 | 0.31 |
| LF6 | Verb + event + prep + wearable + AF<br>- “Got notification from wearable of AF” | 0.67 | 0.02 | 0.20 |
| LF7 | Event + prep + wearable + AF<br>- “Notified on wearable of AF” | 0.74 | 0.10 | 0.27 |
| LF8 | Wearable + subject + verb + AF<br>- “Per wearable, patient had AF” | **0.96** | 0.22 | 0.38 |
| Label Model |  | 0.84 | **0.72** | **0.77** |

*Averages taken from 10-fold cross validation on 600 manually labeled test set. Bold numbers indicate the best observed performance for each metric. LF = Labeling Function. Prep = Preposition. Precision = true positive / (true positive + false positive). Recall = true positive / (true positive + false negative). F1 = 2 \* precision \* recall / (precision + recall).*

Since each labeling function is geared towards identifying positive samples that follow a specific pattern, each labeling function exhibits precision that is significantly higher than recall. By combining these labeling functions into one generative label model, we were able to improve recall. Running the label model on the 56,924 clinical notes flagged 5,829 notes as positive samples, compared to the 105 positive samples in the test set identified through manual labeling.

### Label Model Performance 2: Prompt-Based Labeling Functions

In a day, three prompt-based labeling functions were developed. They were structured so that the notes are presented to the LLM in a zero-shot fashion. Identical prompts were then presented to both LLMs: T0++ and FLAN-T5-XXL. Table 2 shows the performance of each labeling function and the resulting label model.

**Table 2.**
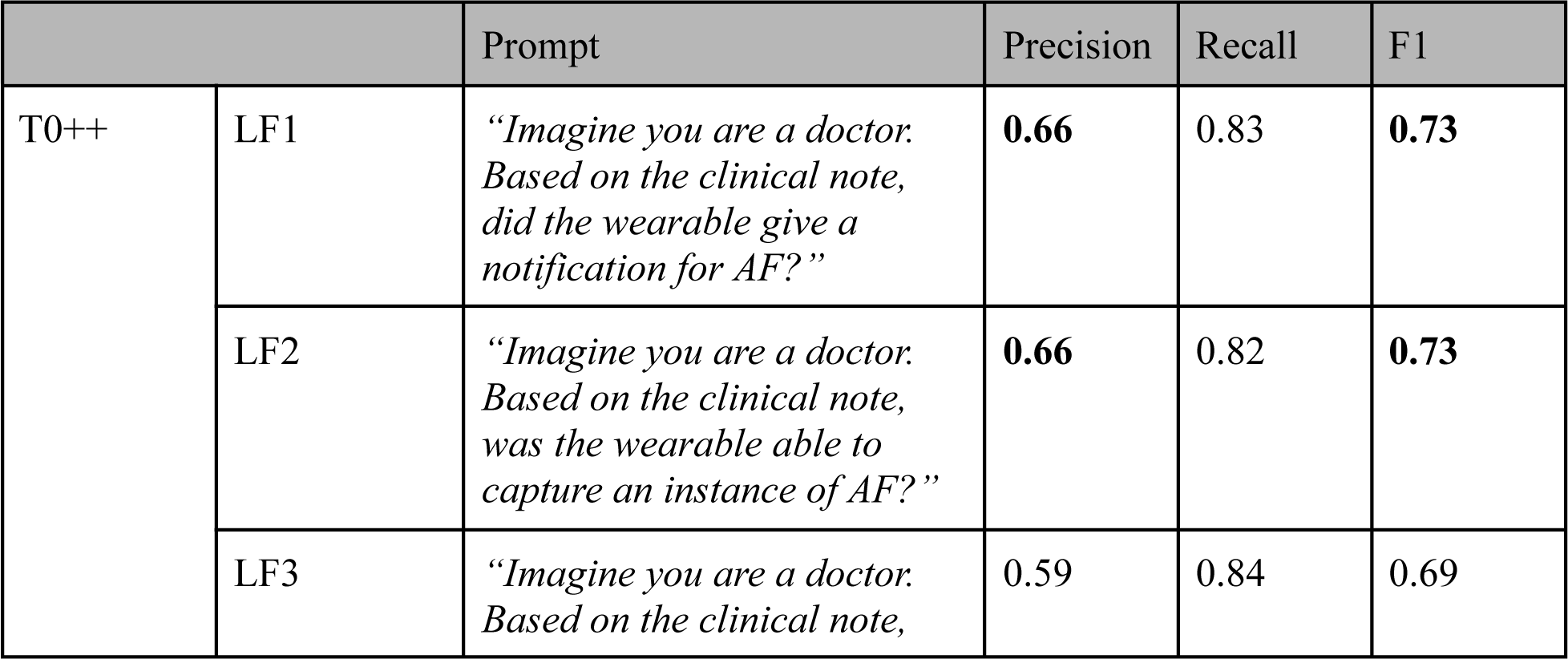
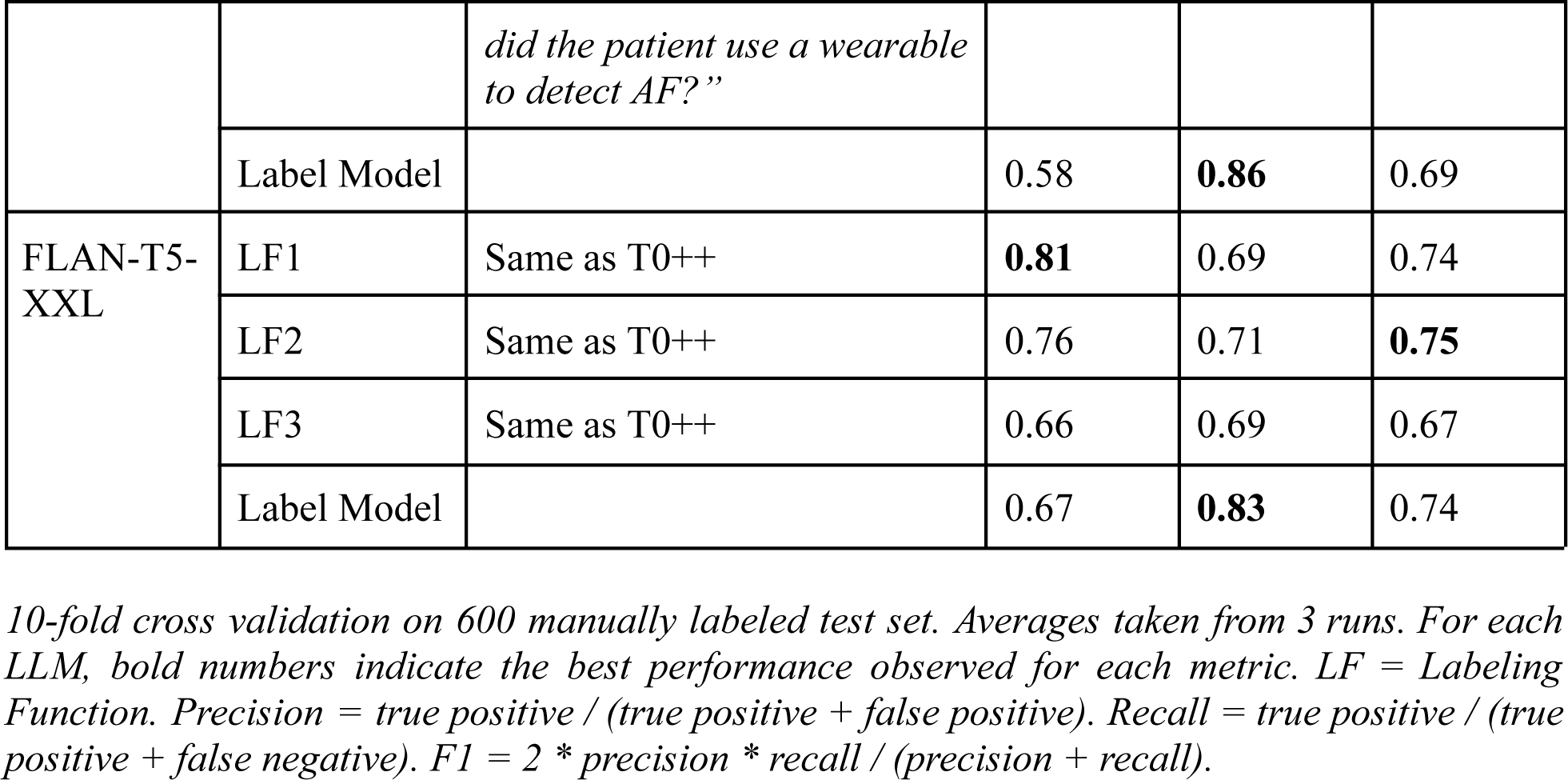
Label model performance with prompt-based labeling functions.

It can be seen that the prompt-based approach, which was developed in one day, manages to attain a comparable portion of the performance achieved through a code-based approach, which was developed with an unbounded amount of time. Specifically, the label model derived by using FLAN-T5-XXL captures ∼96% performance of the label model derived from code-based labeling functions (see Table 1).

Unlike code-based labeling functions, however, prompt-based labeling functions already achieve high recall, due in part to the generalizability of the underlying LLM. Due to this, combining multiple labeling functions with overlapping recall results in interference, and usually ends up in a worse F1 score. In such a case, a single, best-performing label function could be used as a label model instead (a.k.a., distillation).

### Classifier Performance

Since the label model derived from code-based labeling functions exhibited better absolute performance (see Table 1) over prompt-based (see Table 2), we report the performance of the classifier that was fine-tuned using the clinical notes labeled by the label model derived from code-based labeling functions. Table 3 shows the average performance of the classifier on the test set, across varying training set sizes. Training set size was capped at 15,000, since the label model had labeled 5,829 notes as positives. Regardless of the training set size, the test set was held out from the input to the fine-tuning process.

**Table 3.** Classifier performance across varying training set sizes.

| Training Set | Precision | Recall | F1 |
| --- | --- | --- | --- |
| 600 | 0.37 | 0.68 | 0.48 |
| 5,000 | 0.79 | **0.85** | 0.81 |
| 10,000 | 0.84 | 0.81 | **0.83** |
| 15,000 | **0.85** | 0.81 | **0.83** |

*For each training set, average values are reported across 3 runs with different random seeds. For each run, the best performing classifier snapshot was used. Bold numbers indicate the best performance observed for each metric. Precision = true positive / (true positive + false positive). Recall = true positive / (true positive + false negative). F1 = 2 \* precision \* recall / (precision + recall).*

Table 3 readily demonstrates how end model performance benefits from the weakly supervised approach. In particular, input data set size 600 emulates the hypothetical scenario where the size of the training set is limited due to manual labeling overhead. Such a small dataset is not enough to adequately fine-tune a large NLP classifier such as Clinical-Longformer (F1 = 0.48).

As training set size is increased, the classifier obtains better performance, reaching the best average F1 score of 0.83 (precision = 0.85, recall = 0.81). When compared to the label model in Table 1 (precision = 0.84, recall = 0.72), the classifier significantly improves recall, demonstrating that the classifier manages to generalize beyond those rules specified by the labeling functions.

Figure 2 then compares the best performing classifier from each training set size. Again, ROC curve (Figure 2, left) shows that even the best classifier with training set size of 600 performs worse than classifiers from larger dataset sizes. Specifically, in the precision-recall curve (Figure 2, right), the classifier loses significant precision for small gains in recall, further hinting that the classifier is not properly trained. In contrast, comparing the precision-recall curve of larger datasets shows precision increasing with larger training set sizes.

**Figure 2.**
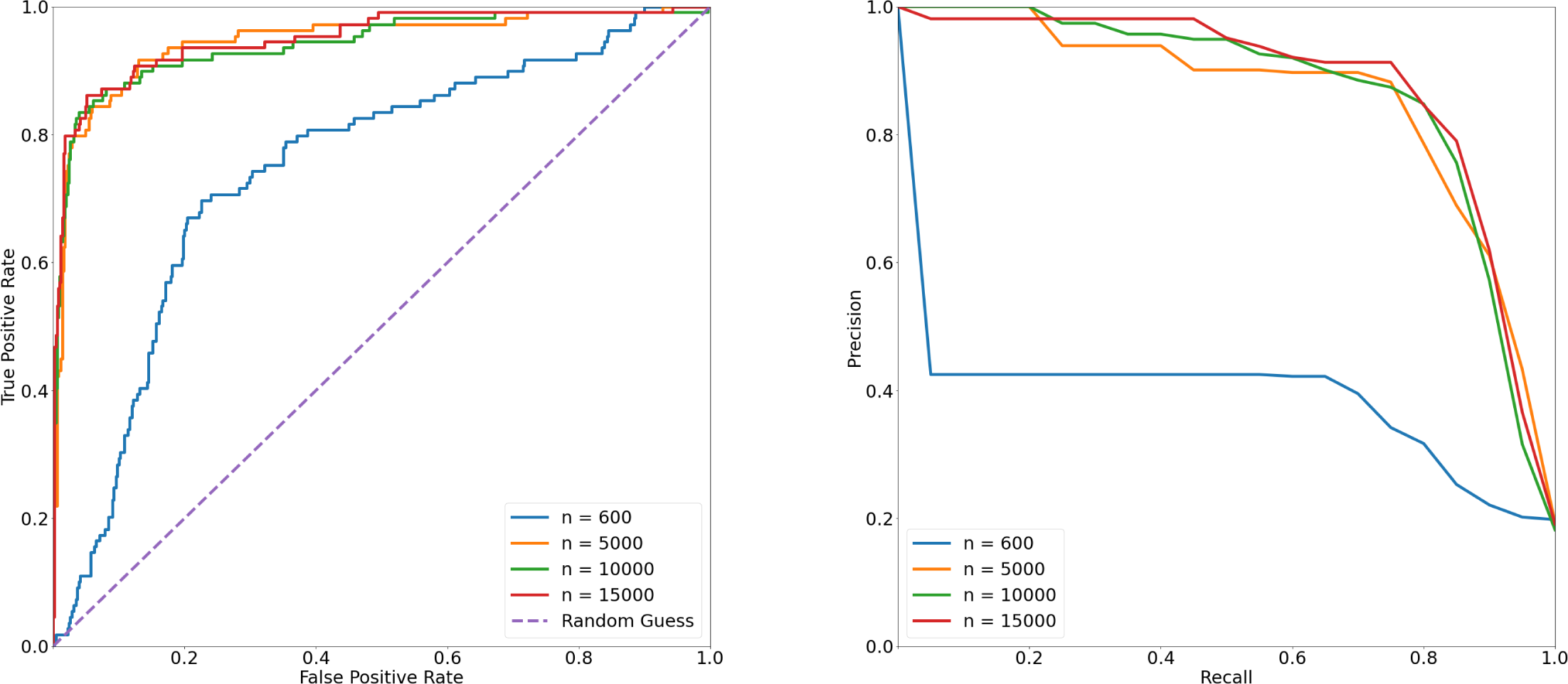
Best performing classifiers across different training set sizes. For each training set, the best performing run was chosen among 3 runs with different random seeds. For each run, the best performing classifier snapshot was chosen.

Across all training set sizes and runs, the best performing classifier achieved F1 = 0.85. Running this classifier on 56,924 clinical notes identified 6,515 notes as containing AF pre-diagnosis events, across 2,279 unique patients.

### Characteristics of Patients

Table 4 compares the summary characteristics of those patients who received AF pre-diagnosis from a wearable device against those who did not. All the patients in the cohort were included, regardless of their prior AF diagnosis.

**Table 4.**
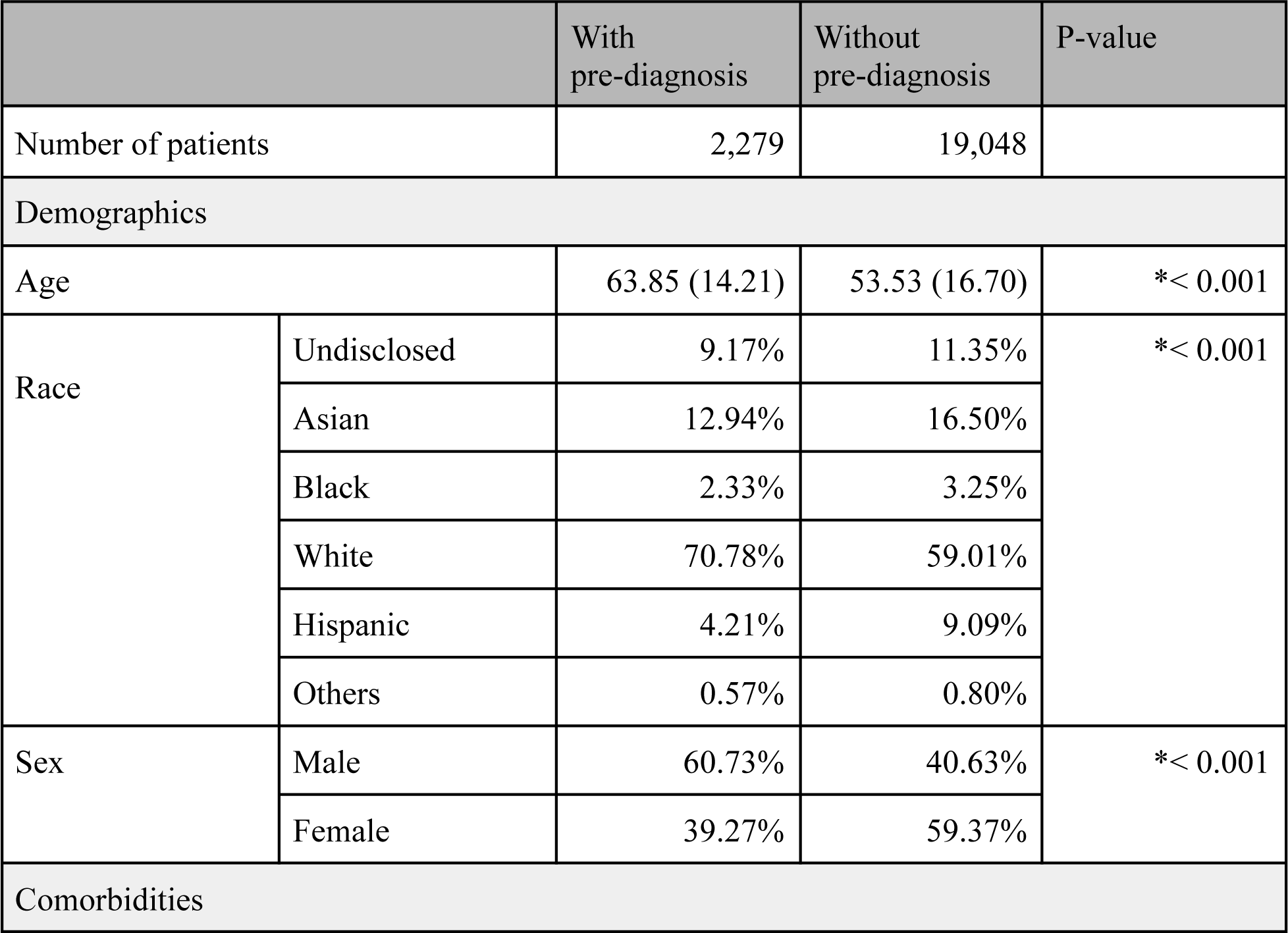
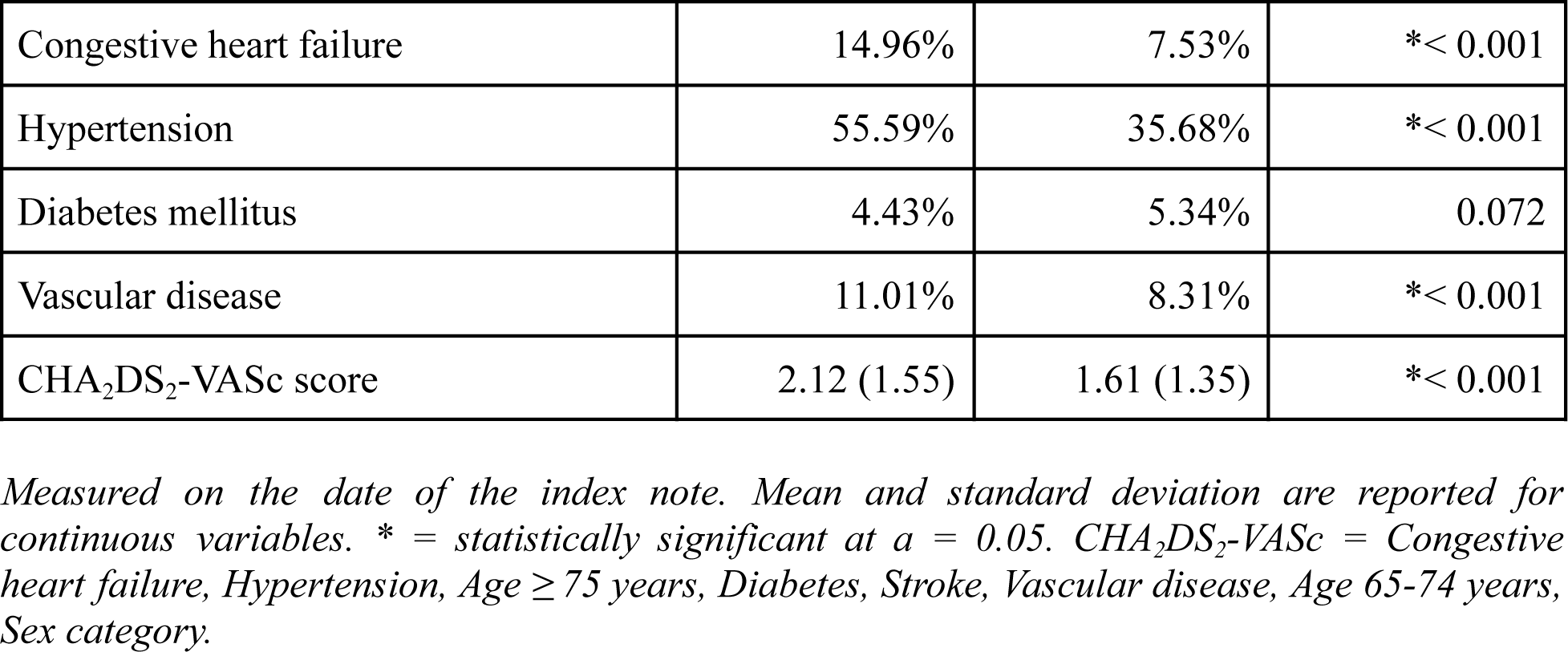
Characteristics of patients.

|  |  | With pre-diagnosis | Without pre-diagnosis | P-value |
| --- | --- | --- | --- | --- |
| Number of patients |  | 2,279 | 19,048 |  |
| Demographics |  |  |  |  |
| Age | | 63.85 (14.21) | 53.53 (16.70) | *$< 0.001$ |
| Race | Undisclosed | 9.17% | 11.35% | *$< 0.001$ |
|  | Asian | 12.94% | 16.50% |  |
|  | Black | 2.33% | 3.25% |  |
|  | White | 70.78% | 59.01% |  |
|  | Hispanic | 4.21% | 9.09% |  |
|  | Others | 0.57% | 0.80% |  |
| Sex | Male | 60.73% | 40.63% | *$< 0.001$ |
|  | Female | 39.27% | 59.37% |  |
| Comorbidities |  |  |  |  |

|  |  |  |  |
| --- | --- | --- | --- |
| Congestive heart failure | 14.96% | 7.53% | * < 0.001 |
| Hypertension | 55.59% | 35.68% | * < 0.001 |
| Diabetes mellitus | 4.43% | 5.34% | 0.072 |
| Vascular disease | 11.01% | 8.31% | * < 0.001 |
| CHA<sub>2</sub>DS<sub>2</sub>-VASc score | 2.12 (1.55) | 1.61 (1.35) | * < 0.001 |

*Measured on the date of the index note. Mean and standard deviation are reported for continuous variables. \* = statistically significant at $\alpha = 0.05$ . CHA<sub>2</sub>DS<sub>2</sub>-VASc = Congestive heart failure, Hypertension, Age $\geq 75$ years, Diabetes, Stroke, Vascular disease, Age 65-74 years, Sex category.*

Patients who received AF pre-diagnosis tend to be older, with more comorbidities except for diabetes mellitus. Race and sex composition are different, with White and male individuals assuming a larger portion of patients who received pre-diagnosis from a wearable. Patients who received pre-diagnosis also exhibit higher CHA_2_DS_2_-VASc scores.

### Characteristics of Patients Without Prior AF Diagnosis

Table 5 then compares the characteristics of patients who had no AF diagnosis prior to index note.

**Table 5.** Characteristics of patients without prior AF diagnosis.

|  |  | With pre-diagnosis | Without pre-diagnosis | P-value |
| --- | --- | --- | --- | --- |
| Number of patients |  | 1,090 | 16,628 |  |
| Demographics |  |  |  |  |
| Age |  | 60.24 (15.64) | 51.56 (16.28) | * < 0.001 |
|  | Undisclosed | 10.37% | 11.94% | * < 0.001 |
| Race | Asian | 12.94% | 17.36% |  |
|  | Black | 2.48% | 3.35% |  |
|  | White | 69.17% | 56.86% |  |
|  | Hispanic | 4.77% | 9.66% |  |
|  | Others | 0.28% | 0.83% |  |
| Sex | Male | 58.90% | 37.72% | *$< 0.001$ |
|  | Female | 41.10% | 62.28% |  |
| Comorbidities |  |  |  |  |
| Congestive heart failure | | 6.79% | 4.13% | *$< 0.001$ |
| Hypertension | | 41.65% | 30.62% | *$< 0.001$ |
| Diabetes mellitus |  | 3.76% | 4.82% | 0.130 |
| Vascular disease |  | 8.07% | 6.50% | *0.049 |
| CHA<sub>2</sub>DS<sub>2</sub>-VASc score | | 1.73 (1.44) | 1.45 (1.23) | *$< 0.001$ |

*Measured on the date of the index note. Mean and standard deviation are reported for continuous variables. \* = statistically significant at $\alpha = 0.05$ . CHA<sub>2</sub>DS<sub>2</sub>-VASc = Congestive heart failure, Hypertension, Age $\geq 75$ years, Diabetes, Stroke, Vascular disease, Age 65-74 years, Sex category.*

Patients without prior AF diagnosis exhibit similar characteristics to the overall cohort, where those who received AF pre-diagnosis tend to be older, White, and male, with more comorbidities except for diabetes mellitus. In particular, 48.07% of the patients who received pre-diagnosis had CHA_2_DS_2_-VASc score 2 or higher, warranting anticoagulation therapy[27]. In contrast, 35.81% of the patients who had not received pre-diagnosis had CHA_2_DS_2_-VASc score 2 or higher.

### Characteristics of Patients with Clinician AF Diagnosis

Among those patients who received pre-diagnosis from a wearable, 12.55% received a clinician AF diagnosis within 60 days from the index pre-diagnosis. Average duration from pre-diagnosis to diagnosis was 7.65 days. Among those patients who never received pre-diagnosis, 1.34% received AF diagnosis within 60 days from the index note.

Table 6 compares clinical characteristics of those patients who received AF diagnosis, based on whether they had received AF pre-diagnosis prior to the diagnosis.

None of the patient characteristics reported in Table 6 differ significantly between those with AF pre-diagnosis and those without, except for the racial composition (p-value = 0.018) and congestive heart failure (p-value = 0.043).

Despite similarities in clinical characteristics, however, patient prescriptions differed based on AF pre-diagnosis. Table 6 shows that those patients with pre-diagnosis tend to be prescribed more with DOAC, with apixaban being the most prescribed.

**Table 6.**
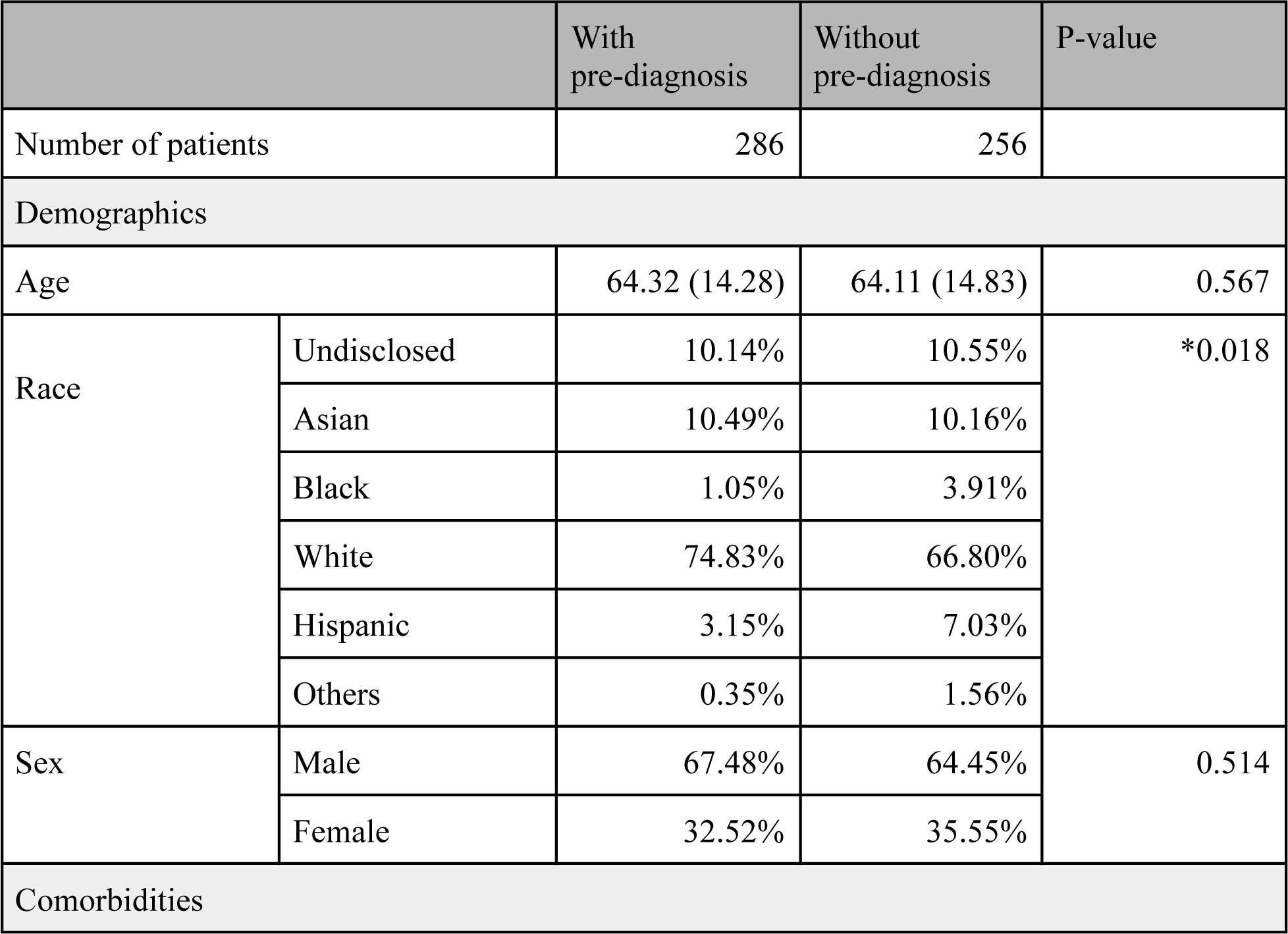
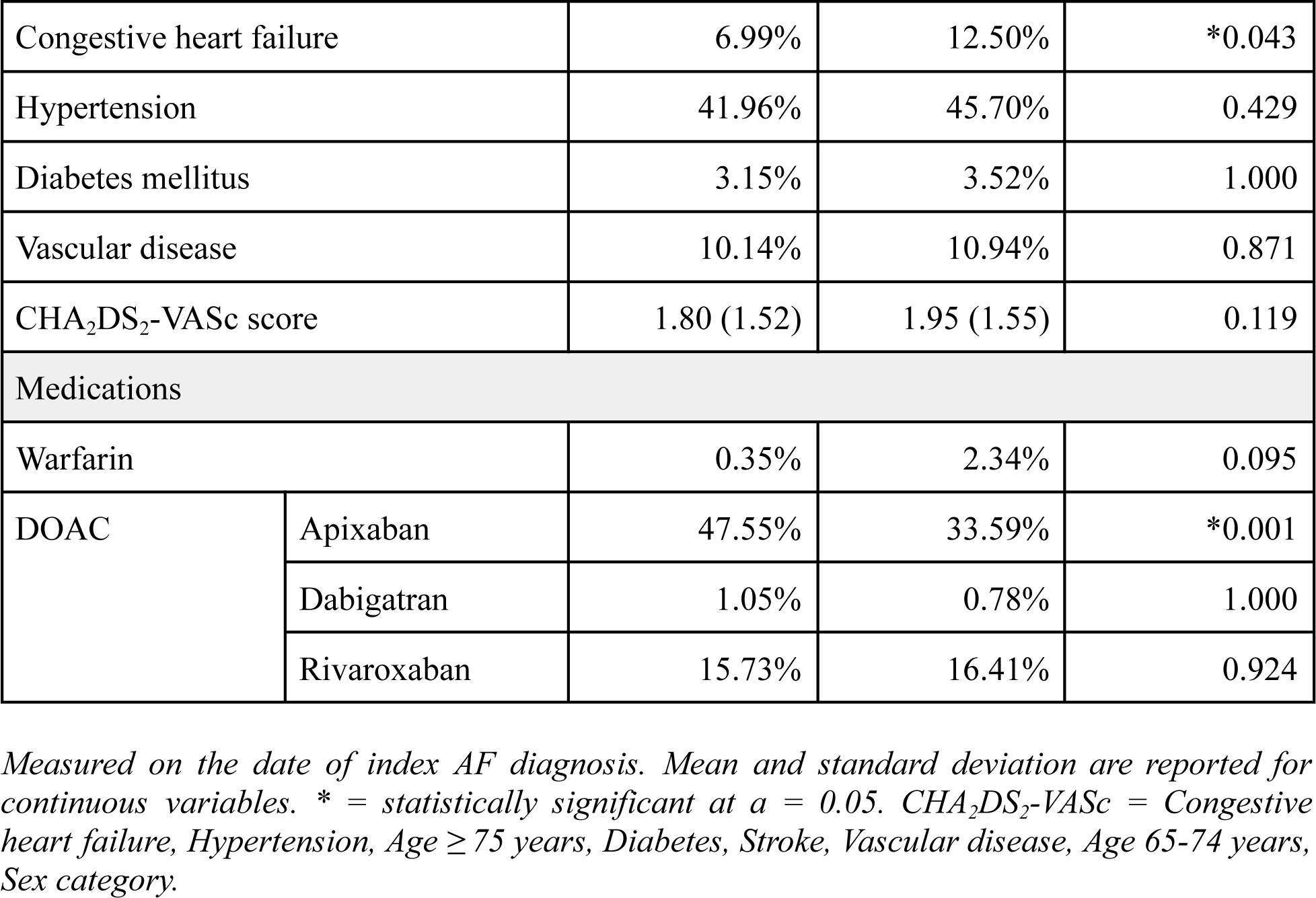
Characteristics of patients with clinician AF diagnosis.

|  |  | With<br>pre-diagnosis | Without<br>pre-diagnosis | P-value |
| --- | --- | --- | --- | --- |
| Number of patients |  | 286 | 256 |  |
| Demographics |  |  |  |  |
| Age |  | 64.32 (14.28) | 64.11 (14.83) | 0.567 |
| Race | Undisclosed | 10.14% | 10.55% | *0.018 |
|  | Asian | 10.49% | 10.16% |  |
|  | Black | 1.05% | 3.91% |  |
|  | White | 74.83% | 66.80% |  |
|  | Hispanic | 3.15% | 7.03% |  |
|  | Others | 0.35% | 1.56% |  |
| Sex | Male | 67.48% | 64.45% | 0.514 |
|  | Female | 32.52% | 35.55% |  |
| Comorbidities |  |  |  |  |
| Congestive heart failure |  | 6.99% | 12.50% | *0.043 |
| Hypertension |  | 41.96% | 45.70% | 0.429 |
| Diabetes mellitus |  | 3.15% | 3.52% | 1.000 |
| Vascular disease |  | 10.14% | 10.94% | 0.871 |
| CHA<sub>2</sub>DS<sub>2</sub>-VASc score |  | 1.80 (1.52) | 1.95 (1.55) | 0.119 |
| Medications |  |  |  |  |
| Warfarin |  | 0.35% | 2.34% | 0.095 |
| DOAC | Apixaban | 47.55% | 33.59% | *0.001 |
|  | Dabigatran | 1.05% | 0.78% | 1.000 |
|  | Rivaroxaban | 15.73% | 16.41% | 0.924 |

*Measured on the date of index AF diagnosis. Mean and standard deviation are reported for continuous variables. \* = statistically significant at $\alpha = 0.05$ . CHA<sub>2</sub>DS<sub>2</sub>-VASc = Congestive heart failure, Hypertension, Age $\geq 75$ years, Diabetes, Stroke, Vascular disease, Age 65-74 years, Sex category.*

## DISCUSSION

In this study we apply weak supervision-based approaches to demonstrate feasibility of developing a post-market surveillance system for medical wearable devices that render AF pre-diagnosis. We first compared two approaches to note labeling, and demonstrated that a label model derived from prompt-based labeling functions using LLMs (precision = 0.67, recall = 0.83, F1 = 0.74) nearly achieved the performance of code-based approach (precision = 0.84, recall = 0.72, F1 = 0.77). We then fine-tuned a classifier on label model output, to accurately identify AF pre-diagnosis (precision = 0.85, recall = 0.81, F1 = 0.83).

Further, by linking the classifier output with structured data in the EHR, we identified patients who were notified via wearables, and conducted a retrospective analysis to compare the baseline characteristics and subsequent clinical treatment of these patients against those who did not receive AF pre-diagnosis. Across the cohort, patients who received pre-diagnosis from a wearable are older with more comorbidities. Race and sex composition of these patients also differ from those who did not receive pre-diagnosis.

We identified a subgroup of patients without prior AF diagnosis, where a higher percentage of patients who received AF pre-diagnosis (48.07% vs. 35.81%) exhibited CHA_2_DS_2_-VASc scores that satisfy anticoagulation recommendation[27]. This enrichment for anticoagulation could be attributed to early pre-diagnosis from the wearable device.

Patients who received pre-diagnosis were 9.37x more likely to receive a clinician assigned AF diagnosis than those who did not. Existence of pre-diagnosis was not correlated with patient demographics or comorbidities at the time of index AF diagnosis, but did correlate with anticoagulation prescription, where patients with AF pre-diagnosis were more frequently prescribed apixaban.

Given that more wearables will be introduced with increasing pre-diagnostic capabilities, and that they have the potential to affect downstream healthcare[3,4], a surveillance framework for wearable devices is urgently needed. However, publications sponsored by wearable vendors so far have focused mostly on ascertaining the accuracy of the pre-diagnostic algorithm itself[1,2]. On the other hand, those publications that sought to conduct post-market surveillance relied solely on manual chart review[3,4], which is hard to scale. In a prior study on wearable notifications, clinician chart review of 534 notified patients yielded only 41 patients with AF pre-diagnosis[3]. In our study, with a weakly-supervised approach, effort spent on clinician review of notes from 600 patients (i.e., the test set) allowed subsequent identification of 2,279 patients with pre-diagnosis.

Approaches that apply various methods of weakly supervised learning to some form of medical surveillance[15,17,28–30] already exist. Most relevant to our work, Callahan et al.[28] apply a weakly supervised approach to implement a surveillance framework for hip implants, and Sanyal et al.[30] for insulin pumps. To the best of our knowledge, however, our work is the first to apply a weakly supervised method using prompting to reduce labeling function authoring burden.

Our results show that the prompt-based approach holds significant potential. While achieving similar performance, the prompt-based labeling functions required only one day of data scientist time, compared to two weeks of data scientist time and three days of clinician time for code-based. In addition, comparing the performance of the label model derived using FLAN-T5-XXL against that of T0++ in Table 2 shows that the same prompt can achieve better performance simply by swapping in a newer, better performing LLM. However, we expect that some extraction tasks are better suited for code-based approaches (e.g., simple dictionary lookup), thus combining code-based and prompt-based approaches is a natural next step.

### Limitations

We acknowledge that the STARR dataset[13] is confined to only two hospitals in a small health care system in a single geographic region. However, our methodology of using a weakly supervised approach to develop a surveillance framework could be readily applied to other institutions. In fact, work is already underway to adapt this approach for use at the Palo Alto VA.

Our results demonstrate that patients who are older, with more comorbidities, White, and male have higher likelihood of receiving AF pre-diagnosis from a wearable. Causality between pre-diagnosis and patient characteristics, however, could not be established. It may very well be that the specific subgroup tends to be health conscious and use wearables more frequently.

## CONCLUSION

By providing pre-diagnosis, medical wearables have the potential to affect subsequent diagnosis and the level of care in the healthcare delivery setting. Post-market surveillance of wearables is necessary to understand the impact of wearables on patient outcomes and health care utilization, but is hindered by the lack of codified terms in EHR to capture wearable use. By applying a weakly-supervised methodology to efficiently detect wearable device AF pre-diagnosis mentions from unstructured EHR data, we demonstrate that such a surveillance system could be built.

Analyzing the AF pre-diagnosis cases in our cohort shows that those patients who received pre-diagnosis exhibit different demographic and comorbidity characteristics from those who did not. We also find that pre-diagnosis from a wearable indeed enriches for anticoagulation and eventual diagnosis of AF. At the index diagnosis, existence of pre-diagnosis from a wearable did not stratify patients on clinical characteristics, but did correlate with the patient anticoagulant prescription.

Our work establishes the feasibility of implementing an EHR-based surveillance system for wearable devices. Further work is necessary to generalize these findings for patient populations at other sites.

## Data Availability

All data analyzed in the present study are available upon reasonable request at https://starr.stanford.edu

## ACKNOWLEDGEMENTS

The authors thank Michael Wornow and Rahul Thapa for their help and support in bringing up LLMs for prompting experiments.

