## Supplemental Material 1 for "Scalable Approach to Medical Wearable Post-Market Surveillance"

### Wearable Surveillance Project Labeling Guideline


**Overview** **1**
**Label List** **2**
**Labeling Schema Descriptions** **2**
Identifying Information 2
note_id and person_id 2
Device Information 3
Device Present 3
Device Version 4
Detection 4
Automatic Notification/Detection 4
On Demand Measurement 6
Irregular Heart Rhythm (Afib) Detection 7
High HR Detected 8
Low HR Detected 9
Fall Detected 11
Oxygen Desaturation Detected 11
Use of device labels 12
Monitoring Irregular Rhythm/AFib 12
Monitoring HR 13
Monitoring Falls 14
Monitoring Wellness 14
Monitoring general health or other 15
Misc Labels 16
Patient Recommended or discussed getting mentioned device 16
Physician used device to render diagnosis 16
**What Constitutes an 'Automatic' Alert** **17**
Wearable 17
Action 17
Event 18
**Canonical Patterns** **19**
Pattern 1: "AFib Noted On Apple Watch" 19
Pattern 2: "Apple Watch Noted AFib" 19
Pattern 3: "Pt Saw Apple Watch Showing AFib" 20
Pattern 4: "Pt Received AFib from Apple Watch" 20
Inversions 20

**Variations on the Pattern** **20**
Event Clarified in Later Sentence 20
Implied Context 21
**Corner Cases** **21**
Recap of Past Encounter 22
Post-Treatment Improvements 22
On-Demand Measurements 22

### Overview

As part of the wearable surveillance project, we are developing a machine-learning based classifier that, given a clinical note, is able to flag whether a patient has received a notification from a wearable regarding the patient's health status. This classifier should also be able to tell which condition/issue the notification was about.

As with any machine-learning model, training this model requires a labeled, ground-truth collection of clinical notes. A subset of these notes are then used to train the classifier, and the rest are used as the answer sheet to quantify model performance.

This document provides a guideline as to how a clinical note should be labeled, to join this ground-truth collection of notes. Through labeling, we apply a set of criteria necessary to determine whether a wearable generated a notification, and which notification it was. If a notification of interest was generated by the wearable, we call the note a 'match', and label it as 'True'. Otherwise the note is labeled as 'False'.

Below we first stipulate how the match result should be documented, then describe the criteria that constitutes a match, provide a few example patterns, discuss some corner cases.

### Label List

| Label Category | **Label Name** | Data Type |
|---|---|---|
| Identifying Information | **note_id** | String |
| Identifying Information | **person_id** | String |
| Device Information | **Device Present** | String |

| | | |
|---|---|---|
| Device Information | **Device Version** | String |
| Alert Information | **Automatic Notification** | Boolean |
| Alert Information | **On Demand Measurement** | Boolean |
| Detection Event | **Irregular Heart Rhythm (Afib) Detection** | Boolean or Blank |
| Detection Event | **High HR Detected** | Boolean or Blank |
| Detection Event | **Low HR Detected** | Boolean or Blank |
| Detection Event | **Fall Detected** | Boolean |
| Detection Event | **Ox Desaturation** | Boolean |
| Use of device | **Monitoring Irregular Rhythm/AFib** | Boolean |
| Use of device | **Monitoring HR** | Boolean |
| Use of device | **Monitoring Falls** | Boolean |
| Use of device | **Monitoring Wellness** | Boolean |
| Use of device | **Monitoring general health or other** | Boolean |
| Misc | **Patient Recommended or discussed getting mentioned device** | Boolean |
| Misc | **Physician used device to render diagnosis** | Boolean |

### Labeling Schema Descriptions

#### Identifying Information

Return to label list (top)

##### note_id and person_id

| | | |
|---|---|---|
| Identifying Information | **note_id** | String |

| Identifying Information | **person_id** | String |
|---|---|---|

"note_id" represents the ID of the clinical note as defined in the STARR-OMOP identified dataset. This ID uniquely identifies the clinical note in the system. "Person_id" represents the ID of the person as defined in the STARR-OMOP identified dataset. This ID uniquely identifies the person in the system.

### Device Information

#### Device Present

[Return to label list (top)](#)

| Device Information | **Device Present** | String |
|---|---|---|

"Device Present" is a mutliclass label used to identify devices described in the note, regardless of whether they generated notification of interest or not. This allows to train the classifier so that it can discern notification of interest from other notifications.

This label should be marked as:

**<name of device>:** If a note mentions any of the wearable devices of interest, this field will contain the string of that **device name**.

**"Unknown device"**: If a note mentions a wearable device, but if the name of the device could not be ascertained, this field will contain the string "unknown device".

Below are some examples:

| Label Value | Example | Reasoning |
|---|---|---|
| **Apple Watch** | *"has an **Apple watch series 3**, and he was very regularly, every approximately 2 days, getting irregular rhythm notifications"* | *Apple watch was mentioned* |
| **Apple Watch** | *"Patient sent a transmission, because his heart rate was irregular (patient wears an **Apple watch**)"* | *Apple watch was mentioned* |
| **Unknown device** | *"Patient got a Afib notification from their **wearable**"* | *No specific "**wearable"** was mentioned* |

##### Device Version

[Return to label list (top)](#)

"Device Version" is a mutliclass label used to identify device <u>versions</u> described in the note. The device version is usually not included, but we thought it would be interesting to keep track of.

This label should be marked as:

**<device version>:** If a note mentions any of the wearable devices of interest AND lists it's specific device version, this field will contain the string of that **device version**.

**<blank>:** Otherwise

| Label Value | Example | Reasoning |
|---|---|---|
| **Series 3** | *"has an **Apple watch series 3**, and he was very regularly, every approximately 2 days, getting irregular rhythm notifications"* | *Apple watch version "series 3" was mentioned* |
| **<Blank>** | *"Patient sent a transmission, because his heart rate was irregular (patient wears an **Apple watch**)"* | *No version was mentioned* |
| **<Blank>** | *"Patient got a Afib notification from their **watch**"* | *No version was mentioned* |

#### Detection

##### Automatic Notification/Detection

[Return to label list (top)](#)
The automatic detection label indicates whether a detection event occurred as an "Automatic Alert", a notification from the wearable device from a measurement that was unprompted by the user. <u>*This is the primary label we are interested in.*</u>

**In order for a clinical note to be considered an automatic alert, three *conceptual components* will likely be present(but do not always need to be): *Wearable(the device in question)*, *Action(what the device did in response to event)*, and *Event(the symptom, condition, or occurrence the device detected)*. See** [What Constitutes an "Automatic Alert" notification](#) for a more detailed understanding. If the note does not fit into the "wearable", "Action", "event" formula, it should have strong evidence to support the presence of an automatic alert if labeled **True.**

**True**: If it can be implicitly concluded that a wearable notification or detection event was received as an ‘automatic alert’, regardless of the presence of any on-demand measurement.

**<Blank>**: **If it can not be concluded** from the note text if the wearable notification or detection event was received as an ‘automatic alert’, **or if there was no notification or detection event mentioned**.

| Label Value | Example | Reasoning |
|---|---|---|
| **True** | “One time at night she was trying to get up from bed and fell. She wears an Apple watch which immediately alerted her daughter.” | Falls are only automatically detected. The Event is a fall, the wearable is an Apple Watch, the action is “alerted her daughter” |
| **True** | “Pt called in, her apple watch notified her 7 times in the last week at night that her HR is below 50 lasting for over 10 mins each time.” | The apple watch notified the patient. Here we can explicitly conclude that the alert was an automatic notification. The event was low heart rate, the wearable was an apple watch, and the action was “notified her 7 times” |
| **<Blank>** | *“She is wearing an Apple Watch and demonstrated in clinic that her watch showed sinus rhythm”* | It can likely be concluded that this was either an on-demand measurement or the patient was showing her doctor her device history. Either way: this would not be a automatic alert as there is no automatic ‘action’ of the device. |
| **<Blank>** | *“She has purchased a new Apple watch to record medical data and to use an APP that will help her recognize when she may be at risk of a fall.”* | *No alert event was mentioned. Therefore, there is no automatic detection alert.* |
| **<Blank>** | *“Notes that he always has "strong symptoms" with atrial fibrillation, and also wears an apple watch that shows that he has been in NSR”* | *No alert event was mentioned. Therefore there is no automatic detection alert. (note that this would be classified as a ‘FALSE’ Irregular Rhythm Detection event, but that is a different column).* |

#### On Demand Measurement

[Return to label list (top)]

The on demand measurement label indicates when a note includes information about how the detection was received. Notifications to alert a user that a symptom was detected can either be produced through an automatic screening process OR a user directed on-demand measurement.

**True**: If it can be implicitly concluded that a wearable notification or detection event was received through a user directed 'on-demand' measurement, regardless of the presence of any automatic alert.

**<Blank>**: **If it can not be concluded** from the note text if the wearable notification or detection event was received from an 'on-demand' measurement, **or if there was no notification or detection event mentioned**.

| Label Value | Example | Reasoning |
|---|---|---|
| **True** | *"She is wearing an Apple Watch and demonstrated in clinic that her watch showed sinus rhythm<br>-continue to monitor your weight daily, symptoms, blood pressure and heart rhythm/rate (via Apple Watch)"* | It can be concluded that a patient used the apple watch to demonstrate NSR with the on-demand feature. |
| **<Blank>** | *"would rather have EKG done sooner rather than later, but patient can do single lead EKG on apple watch to see if in sinus rhythm. Daughter states if done, they will upload image to myhealth. I will follow up on Monday to see if EKG done."* | Although the patient is discussing doing an on demand measurement, this note implies that the EKG has not yet been done. Therefore, we can not say that this patient has performed a EKG and the example should not be labeled as True |
| **<Blank>** | *"Notes that he always has "strong symptoms" with atrial fibrillation, and also wears an apple watch that shows that he has been in NSR"* | It can not be concluded from the text if the patient has taken "on-demand" measurements or is simply relying on automatic detection. Additionally, the watch 'showed' the patient was in NSR (normal sinus rhythm), so no detection event occurred. |
| **<Blank>** | *"Pt called in, her apple watch notified her 7 times in the last week at night that her HR is below 50 lasting for over 10 mins each time."* | *This is an example of and "automatic alert". The on-demand field should be blank and the "automatic alert" label should be True.* |

#### Irregular Heart Rhythm (Afib) Detection

Return to label list (top)
"Irregular Rhythm/Afib" Detection label indicates whether a particular note concludes that **irregular heart rhythm or Afib** was detected by the mentioned device.

As a detection label, this label can have three possible values:

**True**: If it can be concluded that irregular heart rhythm or "atrial fibrillation" was detected by the mentioned device from the note text, regardless of the on-demand vs automatic alert nature of the detection.

**False**: If it can be concluded that irregular heart rhythm or "atrial fibrillation" was **not** detected by the mentioned device from the note text, regardless of the on-demand vs automatic alert nature of the possible detection.

<Blank>: If irregular heart rhythm was **not mentioned or if it can not be concluded** from the note text if irregular heart rhythm or "atrial fibrillation" was detected by the device.

Below are some examples:

| Label Value | Example | Reasoning |
|---|---|---|
| **True** | *"The patient reports fatigue in AF and notes irregularity on an Apple watch."* | *Apple watch noted an irregularity, therefore this note should be considered to have a detection event for irregular rhythm* |
| **True** | *"His apple watch told him that he has atrial fibrillation. He doesn't believe it. His heart rate is down in the 40s and 50s. Limb lead EKG demonstrates atrial fibrillation, and 12-lead was performed."* | *Here we can explicitly conclude that the Apple Watch detected Afib. Therefore, the label should be True* |
| **False** | *"She is wearing an Apple Watch and demonstrated in clinic that her watch showed sinus rhythm<br>-continue to monitor your weight daily, symptoms, blood pressure and heart rhythm/rate (via Apple Watch)"* | The note mentions that apple watch showed sinus rhythm, which implies that no afib was detected. Therefore, the label is False. |
| **False** | *"He also shows me on his apple watch that he has p waves before every QRS and is likely in NSR. He also denies having any afib reports on his apple watch."* | *Patient notes there have been no reports of Afib on Apple Watch: therefore, this label is False* |

| | | |
|---|---|---|
| <Blank> | *"Checked his apple watch/phone, noted HR in 200s."* | *A High HR detection is different than a irregular rhythm detection (see high HR detected label). Since we can not conclude any information about irregular rhythm detection from this note, we leave it blank.* |

#### High HR Detected

Return to label list (top)

"High HR Notification Present" is a boolean label that indicates whether a particular note indicates **high heart rate** (tachycardia) was detected by a wearable. Sometimes physicians will write down that the watch detected a heart rate above a certain number, but not explicitly say that this was 'high'.

This label should be marked as:

**True**: If it can be implicitly concluded that high heart rate aka "tachycardia" was detected by the wearable device from the note text.

**False**: If it can be explicitly concluded that high heart rate notification aka "tachycardia" was **not** detected by the wearable device from the note text.

<Blank>: If high heart rate notification aka "tachycardia" was **not mentioned or if it can not be concluded** from the note text if high heart rate aka "tachycardia" was detected by the wearable.

Below are some examples:

| Label Value | Example | Reasoning |
|---|---|---|
| **True** | *"The apple watch has notified pt 4 notification over 100"* | *Apple watch detected a high heart rate* |
| **True** | *"getting notification from her apple watch due to having high HR ranging from 116-122"* | *Apple watch detected a high heart rate* |
| **True** | *"She recently had a 3-4 hour episode of rapid heart beat, HRs ranging 42-175 bpm (noted on her apple watch)."* | *Apple watch detected a high heart rate (as well as a low heart rate). Since the high heart rate and low heart rate detection criteria were both met, both labels should be true* |

| | | |
|---|---|---|
| **False** | *"He has an Apple watch, but that has not caught any signs of high heart rate"* | *Since the note implies that the watch has not had any high heart rate detections, the label should be False* |
| **False** | *"Does not have BP machine at home but has an apple watch and heart rate remains 50s-100bpm<br>HR on apple watch seems well controlled"* | *The note mentions that the HR is within a homeostatic range and appears to be 'well controlled'. Therefore we can conclude that high heart rate (and low heart rate) have not been detected.* |
| <Blank> | *"Yesterday HR was in high 30s for about 10 minutes - had a notification by her Apple Watch. Feeling more fatigued. No palpitations"* | *A high heart rate detection was not mentioned, and the mentioned heart rate range does not meet the requirements to be considered "high". Therefore, we leave as blank.* |

#### Low HR Detected

Return to label list (top)

| | | |
|---|---|---|
| Notification based use of device | Low HR Notification Present | Boolean or Blank |

"Low HR Notification Present" is a boolean label that indicates whether a particular note indicates that **low heart rate** (bradycardia) was detected by the wearable. Sometimes physicians will write down that the watch detected a heart rate below a certain number, but not explicitly say that this was low.

This label should be marked as:

**True**: If it can be concluded from the note text that low heart rate aka "bradycardia" was detected by the wearable device.

**False**: If it can be concluded from the note text that low heart rate aka "bradycardia" was **not** detected by the wearable device.

**<Blank>**: If low heart rate aka "bradycardia" was NOT mentioned or if it can **not be concluded** from the note text if low heart rate aka "bradycardia" was detected by the wearable.

Below are some examples:

| Label Value | Example | Reasoning |
|---|---|---|

| | | |
|---|---|---|
| **True** | *“apple watch sent him 7 notifications that his heart rate dropped to 40 BPM”* | *Apple watch detected a low heart rate, HR is in range we accept as low* |
| **True** | *“HR was in high 30s for about 10 minutes - had a notification by her Apple Watch”* | *Apple Watch detected low heart rate, HR is in range we accept as low* |
| **True** | *“She recently had a 3-4 hour episode of rapid heart beat, HRs ranging 42-175 bpm (noted on her apple watch).”* | *Apple watch detected a low heart rate (as well as a high heart rate). Since the low heart rate and high heart rate detection criteria were both met, both detection event labels should be true* |
| **False** | *“She has an iwatch and heart rate has been stable- there are no notable episodes of bradycardia arrhythmias”* | *Here we can conclude that there have been no low HR detection events on the iwatch.* |
| **False** | *“Does not have BP machine at home but has an apple watch and heart rate remains 50s-100bpm HR on apple watch seems well controlled”* | *No detection event was mentioned, therefore this is False.* |
| **<Blank>** | *“Occasional his fitbit registers elevated HRs >110, He's not particularly aware of them, and doesn't recall any triggers.”* | *A low heart rate detection was not mentioned, and the mentioned heart rate range does not meet the requirements to be considered “low”. Therefore, we leave as blank.* |

#### Fall Detected

[Return to label list (top)]

| Notification based use of device | Fall Notification Present | Boolean or Blank |
|---|---|---|

This label should be marked as:

**True**: If it can be concluded that a fall was detected by a wearable device from the note text.

<Blank>: If it can not be concluded that a fall was detected or no fall was mentioned

| Label | Example | Reasoning |
|---|---|---|

| Value | | |
|---|---|---|
| **True** | *“One time at night she was trying to get up from bed and fell. She wears a Apple watch which immediately alerted her daughter.”* | A fall was detected. |
| **<Blank>** | *“Per patient, she has an Apple watch set up to detect falls and call for help if needed.”* | *Does not mention if any falls have been detected or not* |
| **<Blank>** | *“His apple watch told him that he has atrial fibrillation.”* | *Does not mention if any falls have been detected or not* |

#### Oxygen Desaturation Detected

Return to label list (top)

| Notification based use of device | Oxygen Desaturation Present | Boolean or Blank |
|---|---|---|

This label should be marked as:

**True**: If it can be concluded that oxygen desaturation was detected by a wearable device from the note text.

**False**: If it can be explicitly concluded that oxygen desaturation was **not** detected by the wearable device from the note text.

<Blank>: If it can not be concluded that oxygen desaturation was detected or no oxygen desaturation was mentioned.

| Label Value | Example | Reasoning |
|---|---|---|
| **True** | *“pt reported nocturnal desats on apple watch and continued daytime symptoms.”* | Implies apple watch detected desaturation |
| **False** | *“No desat notifications from Apple Watch”* | *Explicitly implies no desat detection from watch* |
| **<Blank>** | *“His apple watch told him that he has atrial fibrillation.”* | *Does not mention if any* oxygen desaturation |

### Use of device labels

These labels are used to indicate notes where it can be determined that a patient is using a wearable device in a medically relevant way to monitor specific conditions.

#### Monitoring Irregular Rhythm/AFib

[Return to label list (top)](#)

| | | |
|---|---|---|
| Use of device | **Monitoring Irregular Rhythm/AFib** | Boolean |

**True**: If it can be concluded that **a patient** is using (or plans to use) their device to monitor AFib **or** a physician analyzed the device history for Afib. **Note that this means the intended use of a device must be described by a patient, not be the clinician.**

**<Blank>**: If it can **not** be concluded that a patient is using (or plans to use) their device to monitor AFib or a physician analyzed the device history for Afib.

| Label Value | Example | Reasoning |
|---|---|---|
| **True** | *"Permanent atrial fib: Asymptomatic no events on pacemaker interrogation. She wears her Apple Watch and pays close attention to her HR."* | It is likely that this patient is using the Apple Watch to monitor both her HR and atrial fib. Therefore, both monitoring labels for Afib and HR should be true |
| **True** | *"M with HTN, mixed VHD (moderate MR/AI), and persistent atrial fibrillation on Xarelto, who returns for f/u. He is accompanied by his wife. He currently feels well without new complaints. His new apple watch says he has been in SR, but he feels no different in AF vs. SR."* | *We can conclude that this patient is monitoring Afib condition with his apple watch.* |
| **<Blank>** | *"His apple watch told him that he has atrial fibrillation."* | *This statement does not imply that the patient was using the apple watch to monitor afib prior to this detection event.* |

#### Monitoring HR

| | | |
|---|---|---|
| Use of device | **Monitoring HR** | Boolean |

**True**: If it can be concluded that a patient is using (or plans to use) a device to monitor **HR** for medical purposes or a physician analyzed the device history for **HR**. **Note that this means the intended use of a device must be described by a patient, not be the clinician.**

**<Blank>**: If it can **not** be concluded that a patient is using (or plans to use) a device to monitor **HR** for medical purposes **nor** a physician analyzed the device history for **HR**.

| Label Value | Example | Reasoning |
|---|---|---|
| **True** | *"Okay to to stop metoprolol, advised to monitor BP and for SVTs on his Apple Watch"* | The patient is being instructed to monitor "Supraventricular tachycardia (SVT)" on their apple watch. Since SVT relates to HR, the label is True |
| **True** | *"in the mountains she feels her heart beating and the speed is faster. Her pulse may go to 120, generally it is in the 90's. Based on apple watch"* | *We can conclude that this patient is monitoring their HR with their apple watch.* |
| **True** | *"Pt checks HR using Fitbit. Notes that HR remains around 60-70. 101 on 4/26 while washing dishes. Increases to 120 while walking. Pt states they sent device check to Stanford last night."* | *Patient is monitoring HR using Fitbit device for medical purposes.* |
| **<Blank>** | *-still exercises every day -follows fitbit* | *This statement does not imply that the patient was using the fitbit to monitor HR for medical purposes* |

#### Monitoring Falls

[Return to label list (top)]

| Use of device | **Monitoring Falls** | Boolean |
|---|---|---|

**True**: If it can be explicitly concluded that a patient is using (or plans to use) a device to monitor or predict **falls** or that a physician analyzed the device history for **falls**. **Note that this means the intended use of a device must be described by a patient, not be the clinician.**

**<Blank>**: If it can **not** be explicitly concluded that a patient is using (or plans to use) a device to monitor or predict **falls** **nor** that a physician analyzed the device history for **falls.**

| Label Value | Example | Reasoning |
|---|---|---|
| **True** | “Fall Risk Fall Risk Factors: No recent falls. Pt. wears Apple watch with fall detection” | Patient is using device to monitor fall events |
| **True** | *“Per patient, she has an Apple watch set up to detect falls and call for help if needed.”* | Patient is using device to monitor fall events |
| **True** | *“Had two falls about a month ago. Son bought an apple watch to help alert family in case of future falls.”* | Patient is using device to monitor fall events |
| **True** | *She has purchased a new Apple watch to record medical data and to use an APP that will help her recognize when she may be at risk of a fall.* | Patient is using device to monitor fall events |

#### Monitoring Wellness

Return to label list (top)

| Use of device | **Monitoring wellness (fitness or sleep)** | Boolean |
|---|---|---|

**True**: If it can be explicitly concluded that a patient is using (or plans to use) a device to monitor **their wellness (defined as including fitness and sleep etc) or** that the physician analyzed the device history for **wellness. Note that this means the intended use of a device must be described by a patient, not be the clinician.**

**<Blank>**: If it can **not** be explicitly concluded that a patient is using (or plans to use) a device to monitor **their wellness** **nor** that the physician analyzed the device history for **wellness.**

| Label Value | Example | Reasoning |
|---|---|---|
| **True** | *-continue daily walks (goal is to meet activity target on Apple Watch!)* | Patient is using device to monitor fitness |
| **True** | *“Not as much pysical activity as she used to, but agrees to walk around the building and likes to idea of using a Fitbit to track steps. -patient amenable to walking around her building, advised to obtain a Fitbit to track her step count.”* | Although the patient does not yet have the wearable device mentioned, they are interested in monitoring fitness. Therefore, this label should be true and the “Patient Recommended to get |

| | | |
|---|---|---|
| | | mentioned device" label should also be true |

##### Monitoring general health or other

[Return to label list (top)]

**True**: If it can be explicitly concluded that a patient is using (or plans to use) a device to monitor a condition or physician analyzed the device history for a condition, where the condition is not covered by the other categories. **Note that this means the intended use of a device must be described by a patient, not be the clinician.**

**<Blank>**: If it can **not** be explicitly concluded that a patient is using (or plans to use) a device to monitor a condition **nor** a physician analyzed the device history for an condition, where the condition is not covered by the other categories.

| Use of device | **Patient Monitoring other** | Boolean |
|---|---|---|

| Label Value | Example | Reasoning |
|---|---|---|
| **True** | *"he was given fitbit watch and appears to be more in tune with his chest…"* | Patient is monitoring vague chest related symptoms |
| **True** | *"wearing diapers and at night she will change once but not during the day. Has been wearing a Fitbit tracking her activity at night getting up to void 1-3X/night which is still improved for her."* | Patient tracking how often they go to the bathroom at night with FitBit |
| **True** | *"Palpitations As per her apple watch Being monitored by cardiology"* | Palpitations do not fit into the other categories. (This is actually a bad example because palpitations are not detectable by Apple Watch) |

#### Misc Labels

##### Patient Recommended or discussed getting mentioned device

[Return to label list (top)]

A label to indicate whether the patient was recommended to get the **mentioned device** by their physician or discussed getting one.

**True**: If it can be concluded that a patient **does not have the mentioned wearable device,** and was recommended to get one or discussed getting one.

**<Blank>**: If it can **not** be concluded that a patient **does not have the mentioned wearable device,** or was recommended to get one or discussed getting one.

| Label Value | Example | Reasoning |
|---|---|---|
| **True** | *His other questions pertain to a variety of items: does he need an apple watch?* | Patient discussed or recommended to get mentioned device |
| **True** | *Pt wanted to get permission from PCP she can buy the new apple watch series 5 as a form of emergency if she had a fall (similar to "life alert") She states that she confirmed with an apple specialist this is a capability that the new watches can do.* | *Patient discussed or recommended to get mentioned device* |
| **True** | *Talk with the cardiologist about the apple watch* | Patient discussed or recommended to get mentioned device |

##### Physician used device to render diagnosis

Return to label list (top)

This label indicates if a healthcare provider used information from the device that was mentioned.

**True**: If it can be concluded that the provider utilized information from the mentioned device to make a diagnosis.

**<Blank>**: If it can **not** be concluded that the provider utilized information from the mentioned device to make a diagnosis.

### What Constitutes an ‘Automatic’ Alert

In order for a clinical note to be considered an automatic alert, three *conceptual components* must be present: *Wearable*, *Action*, and *Event*.

#### Wearable

This is the stated word that refers to the *wearable* that a patient interacts with. Wearables of interest include:

- Apple Watch
- Samsung Galaxy Watch
- Google Fitbit
- AliveCor Kardia

Other wearables that do not belong to the above list may come up during the analysis. Such instances should be documented and shared with other researchers.

#### Action

This is the word that describes the *action* that the wearable device took, in order to *notify* the patient of an event. Some of the most frequently used words are:

| Word | Example |
|---|---|
| Notify | *"The apple watch has **notified** pt 4 notification over 100"*<br>*"Apple Watch **notified** him of 8 episodes of "atrial fibrillation.""* |
| Show | *"2 times apple watch **showed** irreg heart rate in last year"*<br>*"Her apple watch reading **showed** atrial fibrillation with a rate of 92"* |
| Alarm | *"his apple watch has been **alarming** with low HR notifications"* |

Often, instead of how the wearable notified the patient, the action documents how the event was *generated*:

| Word | Example |
|---|---|
| Record | *"Apple Watch **recorded** four instances of irregular heart beat"*<br>*"His Apple watch still **records** HR as low as 43 bpm"* |
| Register | *"Has apple watch which has **registered** irregular pulse and notification of AF several times"*<br>*"her Apple Watch **registered** that she had an episode of A. fib 3 days ago"* |

When the subject of the sentence is the patient, however, this action is often described from the patient's perspective:

| Word | Example |
|---|---|
| Receive, get, have | *"checked at her apple watch and **received** a notification that he might have A.Fib"*<br>*"has an Apple watch series 3, and he was **getting** irregular rhythm notifications"*<br>*"she has **had** notifications of an irregular pulse on her Apple watch"* |
| Notice, see | *"pt **noticed** AF on Apple Watch"*<br>*"He **saw** atrial fibrillation again on his Apple watch"* |
| Alert | *"He wears an apple watch and generally when he sees his HR go faster than 160 bpm with exercise he is **alerted** to a problem"* |

Note that in some instances, the action of notification or event generation may be implicit, and should be inferred from the context: e.g.,

| Word | Example |
|---|---|
| - | *"Patient sent a transmission, because his heart rate was irregular (patient wears an Apple watch)"* |

### Event

This component describes the medical condition of a patient that triggered notification. Conditions of interest are:

| Condition | Example |
|---|---|
| Atrial fibrillation | *"received 3 notification on his apple watch he was in **A-fib**"* |
| Tachycardia | *"getting multiple notifications from my iWatch about **elevated HR**"* |
| Bradycardia | *"apple watch sent him 7 notifications that his heart rate **dropped to 40 BPM**"* |

The key here is that **a wearable must have conducted some sort of medical diagnosis**. For example, if the condition at hand is AFib, the wearable must have *suggested* that the patient may be undergoing AFib. In this sense, physicians merely *using* a wearable to take an EKG strip, or a patient *looking* at a wearable to check their BPM should not be considered a match.

Note that a single clinical note can document multiple clinical events: e.g., *"she has been getting notifications from her Apple Watch, sometimes for Afib and sometimes for tachycardia".*

Also note that there could be other, non-medical notifications, such as fall notifications or simple calendar notifications. In the scope of this project, these notifications should not be considered a match.

### Canonical Patterns

Preliminary note review shows that based on how the 3 components are grammatically expressed, matching sentences could be (roughly) categorized into 4 patterns.

Below we color code the 3 conceptual components: Wearable, Action, and Event.

#### Pattern 1: “AFib Noted On Apple Watch”

- Variable heart rate with tachycardia-rate 125 and bradycardia rate 45 that he was sent to notification from his apple watch
- episode of a fast, irregular heart rhythm noted on her Apple watch Apple Heart Study application
- Chest congestion/ wheezing/ Afib notification on iwatch
- Episode of A. fib noted on her apple watch 3 days ago
- AF/AFlutter evident from his latitude device and Apple watch

#### Pattern 2: “Apple Watch Noted AFib”

- Apple Watch recorded four instances of irregular heart beat
- Her apple watch reading showed atrial fibrillation with a rate of 92
- has an Apple Watch which reports that he is in a fib
- last year his Apple Watch has given him 3 warnings of potential A. fib
- Apple watch notifications for possible afib
- his apple watch again reported he was in atrial fibrillation

#### Pattern 3: “Pt Saw Apple Watch Showing AFib”

- Patient checked at her apple watch and received a notification to check with his PCP as he might have A.Fib

- has an Apple watch series 3, and he was very regularly, every approximately 2 days, getting irregular rhythm notifications

#### Pattern 4: “Pt Received AFib from Apple Watch”

- noticed AF on Apple Watch

- He saw atrial fibrillation again on his Apple watch

- noted "a-fib" in Apple watch

- Noted "afib" on Apple Watch which concerned her

- has detected this in atrial fibrillation on her Apple Watch

#### Inversions

Of course, there are inversions that document a notification was **not** received. These notes should be marked as False.

- Wears apple watch and no notifications of AF

- Has apple watch and no notifications for atrial fibrillation

- He did buy an apple watch and has used the ECG feature. He has been in sinus rhythm and has had no notifications for a atrial fibrillation

### Variations on the Pattern

Not all phrases in clinical notes fall nicely in the 4 canonical patterns described above. Below we show samples of variations on the pattern.

#### Event Clarified in Later Sentence

In this example, a message from a wearable may be merely referred to as a ‘notification’, which is further clarified in later sentences.

- received notification from her Apple watch that she might have atrial fibrillation

- received 3 notification on his apple watch he was in A-fib
- she received a notification on her Apple Watch that she was currently in AFib
- received a notification on my Apple Watch that during overnight monitoring of my heart rate, it had detected an irregular rhythm suggestive of atrial fibrillation
- he did take several ECGs with his Apple watch and they all were classified as atrial fibrillation

#### Implied Context

It may not be directly clear that the subject which sent a notification was a wearable device. This should be inferred from the context.

- He does monitor his heart rhythm with his apple watch. He has had notifications for atrial fibrillation
- bought an Apple watch and shortly after putting it on she received an irregular pulse notification suggesting that she might have atrial fibrillation
- Atrial fibrillation with rapid ventricle response-newly diagnosed. By description is likely had brief episodes on and off for the past several weeks but more sustained this morning and notified by apple iWatch

### Corner Cases

There are cases which are considered a match, but require different downstream processing.

#### Recap of Past Encounter

In a medical note, a physician may recap a past encounter in which a patient received notification from a wearable. In this case the date when the note was written can be vastly different from the time when the patient received a notification.

This problem can be fixed by the preprocessing pipeline, by parsing out the date that the notification was sent.

#### Post-Treatment Improvements

There could be cases where a (1) past notification from the wearable led to a clinical diagnosis, (2) corrective actions have been taken (e.g., ablation, anti-coagulation), therefore the patient (3) no longer receives a notification.

Such notes should still be considered a match, since it documents the past existence of such notifications. Preprocessing pipeline should parse out the date when the past notification was sent.

#### On-Demand Measurements

In addition to automated notification, some wearables provide a way to take on-demand measurements (e.g., EKG), and process the measurement to determine whether the patient is experiencing a medical event.

Medical conditions discerned through such on-demand measurement should still be considered a ‘notification’, in that the wearable conducted rudimentary medical diagnosis and presented the outcome to a patient.
