## Supplemental Table 1 for "Scalable Approach to Medical Wearable Post-Market Surveillance"

**Supplemental Table 1. Complete list of search terms for wearable devices.**

| |
|---|
| apple watch |
| iwatch |
| applewatch |
| fitbit |
| fit bit |
| fit-bit |
| galaxy watch |
| samsung watch |
| google watch |
| kardia |
| alivecor |
| alive cor |
| wearable |
| smart watch |
| smartwatch |
